# Feasibility Trial Protocol for a Remote Intentional Music Listening Intervention to Support Mental Health in Individuals with Chronic Stroke

**DOI:** 10.1101/2025.08.15.25333806

**Authors:** V. Provias, M.A. Kucukoglu, A. Robinson, S. Yandun-Oyola, R. He, A. Palumbo, A.J. Sihvonen, Y. Shi, M. Malgaroli, H. Schambra, M. Fuentes, P. Ripollés

## Abstract

**Introduction:** Post-stroke depression affects approximately 30% of stroke survivors and is linked to worse functional outcomes, cognitive decline, reduced quality of life, and increased mortality. While early treatment of post-stroke mental health conditions is critical, current pharmacological options offer limited efficacy. Music listening interventions are a promising, low-risk, accessible, and affordable alternative that may enhance recovery through engagement of reward-related brain circuits. However, most music listening studies have focused on the acute stage of stroke, lack objective measures of music engagement, and rarely assess underlying neural mechanisms. To address these gaps, we propose a feasibility study of a remotely delivered music-listening intervention for individuals with chronic stroke, incorporating objective tracking of music exposure and multimodal assessments of mental health, cognitive, neural, and physiological changes.

**Methods and analysis:** We will conduct a parallel group randomized controlled feasibility trial enrolling 60 patients with chronic stroke from a well-characterized stroke registry in New York City. Participants will be randomized to either an intentional music listening (IML) group or an active control group that listens to audiobooks. The study includes a 4-week pre-intervention period during which no treatment is administered; this phase is designed to assess the stability of outcome measures. Following this, participants will engage in 1-hour daily listening sessions over a 4-week intervention period. All listening activity (i.e., track identity, duration, and engagement) will be continuously tracked using custom open-source software, providing a measure of treatment dose. Behavioral outcomes related to mental health will be assessed at baseline, pre-intervention, post-intervention, and 3-month follow-up. Multimodal biomarkers (functional and structural MRI, electrodermal activity, and heart rate) will be collected pre- and post-intervention. The primary objective is to establish feasibility, defined by rates of retention and adherence, treatment fidelity, feasibility, acceptability, and participant burden. Secondary outcomes include recruitment and randomization rates. This trial will provide essential data to inform the design of future large-scale clinical studies of IML for post-stroke mental health recovery.

**Ethics and Dissemination:** The study was approved by New York University’s Institutional Review Board (FY2024-8826). All human participants will provide written informed consent prior to participation and will be adequately compensated for their time. Results will be reported in peer-reviewed journals.

**Trial registration number:** NCT07127159 (ClinicalTrials.gov).

**Article Summary:** *Strengths and limitations of this study:*

- Access to a large, diverse, and well-characterized population of chronic stroke patients from the greater New York City area.
- Use of a digital platform for intervention delivery and collection of experience sampling and digital phenotyping data.
- Use of multi-modal methods, including an objective measure of treatment dose, to assess the effect of the intervention (behavior, physiology, neuroimaging).
- Limited power to assess mental health, cognitive, and brain plasticity effects of the intervention; the primary focus of the study is to determine feasibility.
- Limited generalizability as this is a single-center study.

## Introduction

Stroke is a leading cause of disability world-wide (WHO, 2021). Upper extremity impairment is one of the most common deficits, affecting over 75% of stroke survivors (Lawrence et al., 2001). However, mental health disorders are also highly prevalent following stroke: meta-analyses indicate that approximately 30% of stroke survivors experience post-stroke depression (Hackett et al., 2005; Hackett & Pickles, 2014). Critically, poor post-stroke mental health is associated with increased risk of long-term disability, greater cognitive impairment, reduced quality of life, and higher mortality rates (Jørgensen et al., 2016; Medeiros et al., 2020; Blöchl et al., 2019; Shi et al., 2017).

Despite strong scientific evidence highlighting the importance of early intervention for post-stroke mental health, the prevalence of long-term psychiatric disorders among stroke survivors has not declined at the clinical level (Hackett & Pickles, 2014). Research shows that early treatment of mental health symptoms after stroke can enhance recovery, improve wellbeing, and increase patient survival (Jorge, et al., 2003; Robinson and Jorge, 2016). While pharmacological treatments such as antidepressants may reduce symptoms of post-stroke depression (Robinson & Jorge, 2016; Hackett et al., 2008), their effectiveness remains uncertain (Allida et al., 2020). These limitations underscore the urgent need for complementary interventions that are safe, affordable, easy to implement, and broadly accessible (Edwards et al., 2023).

Animal models of stroke rehabilitation have demonstrated that enriched environments can enhance brain plasticity and facilitate recovery after injury (Vive et al., 2022; Matsumori et al., 2006; Kempermann et al., 2019; Söderström et al., 2009; Johansson & Belichenko, 2002). Music has been proposed as a form of enriched environmental stimulation (Särkämö et al., 2008), and music-based interventions have shown promise in supporting the well-being of individuals with various clinical conditions, including stroke (Sihvonen et al., 2017). These interventions vary in format and the type of engagement they involve, ranging from active participation through music-making (Altenmüller et al., 2009; Ripollés et al., 2016; Rodríguez-Fornells et al., 2012; Schneider et al., 2007; Grau-Sánchez et al., 2018; Raghavan et al., 2016; Palumbo et al., 2022) to receptive engagement through music listening (Baylan et al., 2016; Sihvonen et al., 2020; Särkämö et al., 2008).

Listening to music is widely regarded as one of the most engaging and emotionally rewarding human activities (Juslin & Västfjäll, 2008). People often use music to regulate mood and enhance wellbeing (Sloboda & O’Neill, 2001; Baltazar et al., 2019). Extensive research shows that music can boost positive affect, reduce stress and anxiety, and alleviate depression across the lifespan, including in older adults (Chin & Rickard, 2014; Mas-Herrero et al., 2023; De Witte, 2020; Hays & Minichiello, 2005; Linnemann et al., 2015). Despite evidence of its benefits, the mechanisms through which music listening improves mental health in both clinical and healthy populations remain poorly understood. Clarifying these mechanisms is particularly important in the context of stroke, where the etiology of post-stroke depression and other mental health conditions is still unknown and may involve disruptions to reward-related neural pathways (Robinson & Jorge, 2016; López-Espuela et al., 2020). Several studies have examined the impact of longitudinal music listening interventions on post-stroke mental health (Särkämö et al., 2008, 2014; Sihvonen et al., 2020; Hewitt et al., 2016; Chen et al., 2012; Tsai et al., 2013; Baylan et al., 2020; Le Danseur et al., 2019; Fan et al., 2024; Sumakul et al., 2020; Dayuan et al., 2022). Särkämö et al. (2008) found that listening to self-selected music for one hour daily reduced depressive symptoms compared to standard rehabilitation, with these improvements linked to increased plasticity in reward-related brain regions (Särkämö et al., 2014). However, the positive effects of music listening on post-stroke mental health (Särkämö et al., 2008, 2014; Chen et al., 2012; Le Danseur et al., 2019; Sumakul et al., 2020; Dayuan et al., 2022) have not always been consistently replicated across studies (Sihvonen et al., 2020; Baylan et al., 2020; Hewitt et al., 2016; Fan et al., 2024; Tsai et al., 2013).

A key limitation of previous music listening interventions for stroke is the lack of objective measures of *treatment dose*: how much music patients listen to, what kind of music is used, how patients engage with it (e.g., attentively or passively), and what other activities they may be doing during music listening. Most studies rely on self-reports or diaries, which provide limited and often unreliable information about actual listening behavior (Baylan et al., 2020; Särkämö et al., 2008). Previous work has also focused primarily on the acute stage of stroke (Baylan et al., 2016), neglecting the chronic stage, where deteriorating mental health continues to increase the risk of long-term disability and mortality. Adherence and retention have also been problematic; for example, one study reported a 24% dropout rate and only 22.2% adherence (Hewitt et al., 2016). Finally, few studies have examined underlying neural mechanisms using multimodal imaging biomarkers such as structural and functional MRI (though see Sihvonen et al., 2020). These limitations, particularly the lack of objective data on treatment dose, make it difficult to determine the specific benefits of music listening in stroke rehabilitation and the mechanisms through which they occur. To enable large-scale clinical trials, there is an urgent need for feasibility clinical trials that address these gaps in the design of music listening interventions.

We propose a feasibility study of a remotely delivered music listening intervention for individuals with chronic stroke, incorporating an objective measure of treatment dose. The intervention is guided by the NIH Music-Based Intervention Toolkit (Edwards et al., 2023) and will provide participants with a fully configured iPad equipped with Spotify for music streaming. An open-source tracking software developed for this study will objectively record listening duration and the specific songs played. Using the exact record of music listened to by participants, we will apply music information retrieval (MIR) methods (McFee et al., 2015) to extract the acoustic and musical properties (i.e., the *acoustic fingerprint*) of the music patients engage with on a daily basis. To enhance engagement, adherence, and treatment fidelity, the iPad will also serve as a communication tool for daily checkins and personalized playlist updates. The protocol includes longitudinal assessments—spanning baseline, pre- and post-intervention, and a 3-month follow-up—of behavioral mental health and cognitive outcomes, along with multimodal neural and physiological biomarkers (functional and structural MRI, electrodermal activity, and heart rate). Feasibility will be assessed in a small randomized controlled trial (N=60), including a control group listening to audiobooks, through metrics such as recruitment, randomization, retention and adherence, treatment fidelity, feasibility, participant burden, and acceptability. This study will lay the groundwork for future large-scale clinical trials targeting mental health recovery in chronic stroke.

## Methods and Analysis

### Study Design

We propose a small-sample parallel group randomized controlled feasibility trial where patients will be assigned to an intentional music listening (IML) group (N=30) or to an active control group (N=30) listening to audiobooks (see Figure 1). IML refers to the active practice of listening to music (e.g., paying attention to the music; Dingle et al., 2021). Audiobook listening is a well-established control condition for IML interventions and is recommended by the NIH Music-Based Intervention Toolkit (Edwards et al., 2023; Särkämö et al., 2008; Sihvonen et al., 2020; Baylan et al., 2020). Both groups will complete 1-hour long listening sessions daily for a period of 4 weeks (excluding weekends; 20 sessions total). We will track the music and audiobooks listened to by participants using custom open-source software developed by our team. The study protocol will include a baseline assessment, followed by a pre-intervention period of 4 weeks to assess the stability of outcome measures. Daily control measures will be collected during the pre-intervention period using ecological momentary assessments (EMAs). After the 4-week pre-intervention period, a pre-intervention evaluation session will be completed. The intervention will then start. As in the pre-intervention period, daily control measures will continue to be collected during the intervention using EMAs. After the 4-week intervention period, a post-intervention assessment will be conducted, with a follow-up at 3 months. Baseline, pre-, and post-intervention evaluations will include clinical outcomes (i.e., behavioral tests of mental health and cognition). Pre- and post-intervention evaluations will include biomarkers (i.e., structural and functional MRI and physiology) in addition to clinical outcomes. The follow-up session will only include clinical outcomes. Table 1 provides an overview of all measures collected throughout the study and the time points at which they are administered.

**Table 1.** Summary of data collected.

|  | Screening | Baseline<br>Evaluation | Pre-Inter.<br>Period | Pre-Inter.<br>Evaluation | Intervention | Post-inter.<br>Evaluation | Follow-up<br>Evaluation |
| --- | --- | --- | --- | --- | --- | --- | --- |
| **Eligibility** |  |  |  |  |  |  |  |
| Hearing Handicap Inventory for the Elderly Screening Version (HHIE-S) | X |  |  |  |  |  |  |
| Montreal Cognitive Assessment (MoCA) | X |  |  |  |  |  |  |
| Montreal Battery Evaluation of Amusia (MBEA) | X |  |  |  |  |  |  |
| Barcelona Music Reward Questionnaire (BMRQ) | X |  |  |  |  |  |  |
| MRI Safety Questionnaire | X |  |  | X |  | X |  |
| **Individual Differences** |  |  |  |  |  |  |  |
| Demographics |  | X |  |  |  |  |  |
| Musical/Audiobook preferences |  | X |  |  |  |  |  |
| Goldsmiths Musical Sophistication Index (Gold-MSI) |  | X |  |  |  |  |  |
| **Mental health clinical outcomes** |  |  |  |  |  |  |  |
| Beck Depression Inventory (BDI) |  | X |  | X |  | X | X |
| Stroke-Specific Quality of Life (SS-QOL) |  | X |  | X |  | X | X |
| Perceived Stress Scale (PSS) |  | X |  | X |  | X | X |
| Generalized Anxiety Disorder (GAD-7) |  | X |  | X |  | X | X |
| Positive and Negative Affect Schedule (PANAS) |  | X |  | X |  | X | X |
| World Health Organization Well-Being Index (WHO-5) |  | X |  | X |  | X | X |
| **Cognitive Clinical Outcomes** |  |  |  |  |  |  |  |
| Processing Speed |  | X |  | X |  | X | X |
| Working Memory |  | X |  | X |  | X | X |
| Verbal Memory |  | X |  | X |  | X | X |
| Attention |  | X |  | X |  | X | X |
| **Biomarkers** |  |  |  |  |  |  |  |
| Structural MRI |  |  |  | X |  | X |  |
| Functional MRI |  |  |  | X |  | X |  |
| Physiology |  |  |  | X |  | X |  |
| **EMAs** |  |  |  |  |  |  |  |
| Motor, sensory, cognitive, and social activity |  |  | X |  | X |  |  |
| Music activity outside listening session |  |  | X |  | X |  |  |
| Profile of Mood States (POMS) |  |  | X |  | X |  |  |
| **Feasibility Measures** |  |  |  |  |  |  |  |
| Burden Ratings (Patients) |  | X |  | X |  | X | X |
| Acceptability Ratings (Patients) |  |  |  |  | X |  |  |
| Feasibility Ratings (Team Member) |  |  |  |  | X |  |  |
| Fidelity Ratings (Team Member) |  |  |  |  | X |  |  |

**Figure 1.**
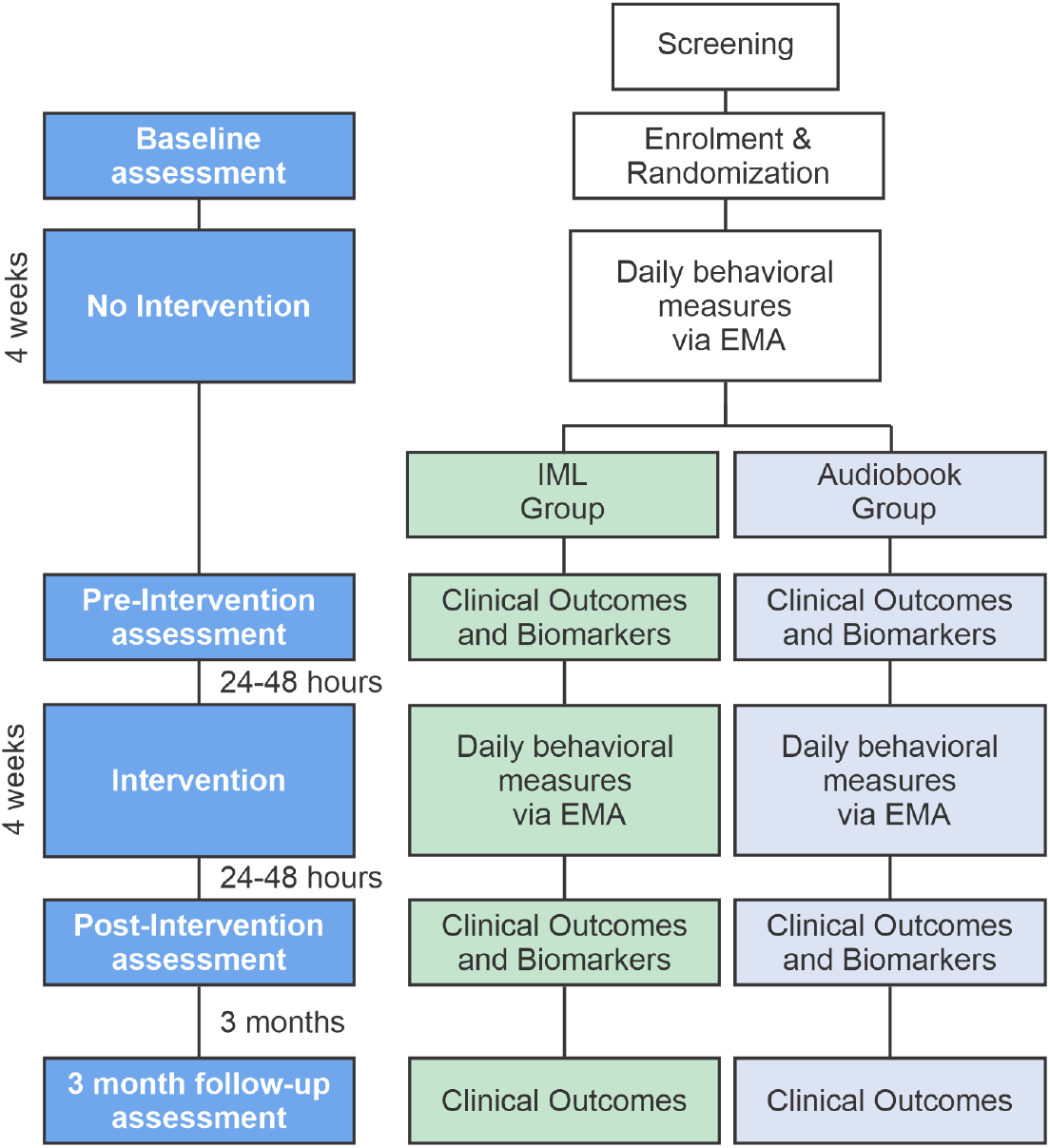
Clinical trial design.

### Study setting

Potentially eligible participants will be identified through a well-characterized registry of stroke patients living in New York City, USA. The patient registry used for recruitment already includes most of the required inclusion and exclusion criteria. Any missing information will be obtained via a follow-up screening session delivered remotely or in person. All study evaluations will be conducted at New York University, while the intervention will be delivered remotely. Research assistants will administer the listening sessions and will not be music therapists.

### Study population

We aim to enroll 60 participants (30 per group), aged 50 to 90, with a confirmed diagnosis of ischemic or hemorrhagic stroke occurring at least six months prior to enrollment. The sample size was informed by previous feasibility studies of complementary health interventions (Richard-Lalonde et al., 2023; Doorley et al., 2022; Portz et al., 2023; Smith et al., 2022). Participants will be excluded if they meet any of the following criteria: (1) significant hearing loss, defined by a score >26 on the Hearing Handicap Inventory for the Elderly Screening (HHIE-S; Ventry and Weinstein, 1982); (2) contraindications for MRI; (3) cognitive impairment that may interfere with completion of study procedures, defined as a Montreal Cognitive Assessment (MoCA; Nasreddine et al., 2005) score below 22 (unless the lower score is attributable to expressive aphasia); (4) specific musical anhedonia, defined as a score below 60 on the Barcelona Music Reward Questionnaire (BMRQ; Mas-Herrero et al., 2013); or (5) amusia, defined as a score below 70% on the Montreal Battery of Evaluation of Amusia (MBEA; Peretz et al., 2003). Participants will not be excluded if they are currently taking medications that may affect brain function (e.g., antidepressants) or if they are engaged in other complementary therapies (e.g., mindfulness, yoga). However, they will be instructed not to initiate any new medications or therapies during the study period. Participants who begin a new treatment during the study will be excluded from the analysis. Participants will also be discontinued from the study upon their request or if there is a worsening of their medical condition that compromises safety or study participation.

Eligible patients will be contacted by phone, and the study will be explained using a standardized script. Patients who do not make an immediate decision (e.g., need time to consider participation) will receive up to two follow-up contacts via phone or email over a two-week period. To meet recruitment targets, we aim to enroll at least five participants per month over the course of 12 months.

### Procedures and assessments

#### Screening

Eligibility will be assessed using a well-characterized stroke registry. Initial screening will verify age and confirm a diagnosis of ischemic or hemorrhagic stroke occurring at least six months prior to enrollment. Additional exclusion criteria will then be reviewed. If data for key screening measures are missing from the registry, a study team member will contact the patient to explain the study and gauge interest. If the patient agrees to participate, they will complete a screening session (online or in person) to confirm eligibility. This session may include: (1) the Hearing Handicap Inventory for the Elderly – Screening version (HHIE-S; Ventry & Weinstein, 1982), a 10-item self-report measure of hearing loss validated against pure-tone audiometry (Lichtenstein et al., 1988); (2) the Montreal Cognitive Assessment (MoCA), a standardized screening tool used to detect cognitive impairment and early signs of dementia (Nasreddine et al., 2005). The MoCA assesses multiple cognitive domains, including executive function (e.g., trail-making), visuospatial abilities (e.g., clock-drawing), attention (e.g., digit span), language (e.g., sentence repetition and verbal fluency), abstraction (e.g., identifying similarities between two concepts), short-term memory (via a delayed recall task), and orientation (e.g., stating the current date, month, and year); (3) the Montreal Battery of Evaluation of Amusia (MBEA; Peretz et al., 2003), assessing the presence of amusia, a disorder affecting pitch and melody perception. Participants hear pairs of short melodies and judge whether they are the same or different, across separate pitch and rhythm discrimination tests; (4) the extended Barcelona Music Reward Questionnaire (eBMRQ; Mas-Herrero et al., 2013; Cardona et al., 2022), a 24-item self-report questionnaire that assesses individual differences in sensitivity to music-related reward. The eBMRQ includes subscales for musical seeking, emotion evocation, mood regulation, social reward, sensory-motor, and music absorption. To screen for specific musical anhedonia, only the original BMRQ score (excluding the music absorption items) will be used; and (5) an MRI safety screening questionnaire assessing eligibility criteria for neuroimaging (see Supplementary Materials). If the patient meets all eligibility criteria, they will be enrolled in the study, randomized to either the IML or control group using a computer-generated randomization sequence, and scheduled for a baseline evaluation session. Experimenters will not be blinded to group allocation.

#### Baseline evaluation session

During the baseline evaluation session, we will first collect measures of *individual differences*, including (1) demographics: age, gender, race, ethnicity, dominant hand, languages spoken, current medications (with dosages), participation in complementary health activities (e.g., yoga, meditation), occupational status, and highest education level (see Supplementary Materials); (2) music/audiobook preferences: participants will report their preferred genres (e.g., music: classical, pop, jazz; audiobooks: science fiction, noir, biography), favorite artists/authors, and mood-based music/audiobook preferences to support individualized playlist creation (see Supplementary Materials). Both groups will also provide a list of songs that elicit chills or intense emotional responses, which will be used as stimuli during fMRI scanning; and (3) the Goldsmiths Musical Sophistication Index (Gold-MSI): a 38-item self-report questionnaire assessing musical expertise across dimensions such as active engagement, perceptual ability, training, singing skill, and emotional responsiveness (Müllensiefen et al., 2014). Next, participants will complete behavioral questionnaires assessing *mental health* and *cognitive functioning*, which serve as the primary clinical outcomes. While the main focus is on the impact of IML on mental health, cognitive measures are included to capture domains previously shown to improve with music listening interventions (Särkämö et al., 2008; Sihvonen et al., 2020).

The *mental health measures* will include: (1) the Beck Depression Inventory (BDI), a 21-item self-report questionnaire designed to assess the severity of a person’s depression symptoms (Beck et al., 1961); (2) the Stroke-Specific Quality of Life (SS-QOL), a 49-item self-report questionnaire designed to assess the health-related quality of life specific to survivors of stroke (Williams et al., 1999). It contains 12 subscales: energy, upper extremity function, work, mood, self-care, social roles, family roles, vision, language, thinking, and personality; (3) the Perceived Stress Scale (PSS), a 10-item questionnaire that assesses the degree to which individuals perceive situations in their lives as stressful (Cohen et al., 1983); (4) the Generalized Anxiety Disorder-7 (GAD-7), a 7-item self-report questionnaire that assesses the severity of generalized anxiety symptoms (Spitzer et al., 2006); (5) the Positive and Negative Affect Schedule (PANAS), a 20-item self-report questionnaire designed to assess positive and negative affect (Watson et al., 1988). The scale consists of two 10-item subscales, each measuring the extent to which individuals have experienced a range of positive (e.g., enthusiastic, inspired) or negative (e.g., upset, nervous) emotions; and (6) the World Health Organization Well-Being Index (WHO-5), a 5-item questionnaire designed to assess the mental well-being of individuals, including aspects of vitality, emotional state, and life satisfaction (Topp et al., 2015).

The *cognitive functioning* measures will be assessed using the NIH Toolbox Cognition Battery (Weintraub et al., 2013). We will assess cognitive functioning across several domains: (1) processing speed, using the Oral Symbol Digit Test, a timed task in which participants match symbols to numbers; (2) working memory, using the List Sorting Working Memory Test, which requires participants to recall and sequence items presented both visually and aurally; (3) verbal memory, using the Rey Auditory Verbal Learning Test, where participants are asked to freely recall as many words as possible from a list of 15 unrelated words; and (4) attention, using the Flanker Inhibitory Control and Attention Task, which evaluates the ability to focus on a central stimulus while inhibiting attention to surrounding distractors.

At the end of the evaluation session, participants will also complete a *burden rating*, indicating whether they found the session “not burdensome,” “partially burdensome,” or “very burdensome.”

#### Pre-intervention period

After the baseline assessment, participants will undergo a 4-week no-intervention period to assess the stability of outcome measures. During this time, they will use the provided iPad to complete daily Ecological Momentary Assessments (EMAs), which will track mental health status and collect additional control measures. EMAs will be administered through the mEMA app (https://ilumivu.com/solutions/ecological-momentary-assessment-app/), with participants receiving daily notifications prompting them to complete a brief survey within a six-hour window (6:00 p.m. to midnight). Two surveys will be administered through the mEMA app during the 4-week pre-intervention period: (1) the first is a daily survey assessing control variables related to the four mechanistic pathways through which enriched environments are thought to promote brain plasticity and recovery: motor, sensory, cognitive, and social stimulation (Vive et al., 2022). This survey uses Likert scales to measure the extent of motor, sensory, cognitive, and social activity throughout the day, as well as music engagement. Specifically, whether participants listened to music outside of scheduled sessions and whether the listening was intentional (see Supplementary Materials). Participants will complete this survey each weekday starting after the baseline session; and (2) the Profile of Mood States (POMS), a 36-item standardized self-report questionnaire that assesses temporary mood states across six subscales: Tension-Anxiety, Depression-Dejection, Anger-Hostility, Vigor-Activity, Fatigue-Inertia, and Confusion-Bewilderment (McNair et al., 1971). The POMS will be administered every three days beginning on the first day of the 4-week period (i.e., days 1, 4, 7, 10, 13, 16, 19, 22, 25 and 28, for a total of 10 data points).

#### Pre-intervention evaluation session

After the 4-week pre-intervention period, participants will complete a pre-intervention evaluation session. In addition to repeating behavioral clinical outcome measures (i.e., mental health and cognitive assessments), this session will include the collection of neural and physiological biomarkers. We will acquire functional and structural MRI data on a 3T Siemens Prisma scanner equipped with a 64-channel phased-array head coil. *Neuroanatomical biomarkers*. We will examine biomarkers related to grey and white matter. For grey matter we will collect a high resolution T1 MPRAGE image (TR = 2400 ms, TE = 2.24 ms, flip angle = 8°, voxel size = 0.80 × 0.80 × 0.80 mm^3^, 256 sagittal slices, acq. matrix = 320 × 300). For white matter, we will collect diffusion-weighted images (DW-MRI; one run with ten interleaved non-DW volumes and 128 DW-ones, b-value=1500 s/mm2, 2×2×2 mm^3^).

##### Neural circuits and function

We will collect fMRI data (sequential whole-brain multi-echo echo-planar imaging volumes; TR = 1500 ms, TE=45 ms, flip angle = 77°, voxel size = 1.5 × 1.5 × 1.5 mm^3^, 64 axial slices, acquisition size = 104×104) while participants listen to self-selected rewarding songs and emotionally neutral music (i.e., elevator music; Abrams et al., 2024) across three runs. Each run will consist of four trials, including two rewarding and two neutral excerpts presented in randomized order. Each excerpt will last 60 seconds, followed by a 30-second rest period. After each excerpt, participants will rate their perceived pleasure and familiarity using a 5-point Likert scale. At the end of the evaluation session, participants will also complete a burden rating.

##### Physiological biomarkers

Physiological data (electrodermal activity and heart rate) will be acquired concurrently with the neural data while participants listen to music using an MRI-compatible Biopac system.

#### Intentional Music Listening Intervention

Patients will engage in daily one-hour listening sessions over a four-week period (excluding weekends; total: 20 sessions). Each participant will be provided with an iPad equipped with a prepaid SIM card, allowing them to stream music freely without relying on personal internet access. The iPad will come preloaded with Spotify (for music streaming and playlist creation), FaceTime (for video communication with team members), and the mEMA app (for ecological momentary assessments). During the baseline session, a team member will familiarize each participant with the iPad and its key features, including FaceTime, Spotify, and the mEMA app. Team members will then work with participants to co-create a playlist of self-selected, culturally relevant songs. This playlist can be updated at any time by the patient independently or with support from the team. To promote flexibility and adherence, team members will have remote access to participants’ Spotify accounts and may assist with updating or playing music based on the participant’s preferences.

Each daily remote listening session will begin with a team member calling the patient on the iPad at a pre-arranged time. At the beginning of the session, the playlist will be updated according to the patient’s preferences and mood. Patients will then be asked to listen intentionally to the playlist without engaging in other activities (e.g., doing chores). However, participants will be allowed to engage in music-related behaviors during listening, such as singing or moving to the music. The total duration of each session will be one hour, including at least 40 minutes of music listening and up to 20 minutes for setup, preparation, and playlist updates. Team members will remotely monitor the listening sessions and will be available via FaceTime throughout the session to provide support if needed. This will improve adherence to the protocol. To accommodate varying levels of impairment, needs, and technological familiarity, team members will provide as much support as each participant requires, while also encouraging patient agency. Participants will be guided as needed but allowed to manage their listening sessions independently (e.g., by skipping songs on their own). Listening sessions will also be video-recorded to allow for later annotation of music-related behaviors (e.g., dancing). If video monitoring or recording fails due to technical issues, the research team will first attempt to resolve the problem remotely via FaceTime. If the issue cannot be resolved remotely, a team member will visit the participant in person to fix the problem or replace the iPad. If feasible, a make-up session will be conducted on the same day. At the end of each session, the monitoring team member will rate the feasibility of the session (“not feasible”, “partially feasible” or “fully feasible”) and the participant will rate the acceptability of the session (“not acceptable”, “partially acceptable” or “fully acceptable”). Finally, a team member who was not involved in the listening session will review the session’s video recording and score its fidelity as either “delivered according to protocol” or “not delivered according to protocol.”

The same control measures collected during the pre-intervention period will continue to be gathered during the intervention using EMAs via the mEMA app (motor, sensory, cognitive, and social activity; music engagement outside of scheduled sessions; and the POMS).

#### Control Intervention

The control group will consist of chronic stroke patients who will intentionally listen to audiobooks instead of music. Assessing a group of patients listening to audiobooks will allow us to control any effects unrelated to music (e.g., intentional listening, non-music specific auditory stimulation, social interaction via the phone calls with the team members, etc.). The control group will follow the same protocol as the IML group, ensuring that both groups are matched for treatment intensity. Audiobooks will be played via Spotify and will be self-selected and culturally relevant to each participant, with the option to update selections during the 4-week intervention period. Audiobooks will not contain background music. Importantly, if participants in the control group typically listen to music as part of their daily routine, they will be asked to do so using the Spotify app on the provided iPad. No curated playlists will be created for the control group. This approach will allow, for the first time in an IML intervention, objective tracking of music use in the control condition. After the follow-up evaluation session, control participants will be informed of the potential benefits of IML.

#### Tracking Software

To track each patient’s music and audiobook listening as objectively and precisely as possible, we developed a Python script that automatically logs the Spotify playback activity of a given account. The script utilizes the Spotify Web API in a while loop, requesting device and playback information from the server in each iteration. After each iteration there is a two-second pause to comply with Spotify rate limits. To track when the Spotify account is opened and closed the script must run continuously, which requires a dedicated computer. We will use the NYU Supercomputer Greene for this purpose. Participants’ music and audiobook listening will be continuously tracked using this custom Python script. Running the script continuously will also allow us to capture music or audiobook listening that occurs outside of the scheduled sessions with equal precision.

The tracking script is initiated by entering the patient’s Spotify account ID, which serves as their unique identifier for monitoring listening activity. It then reads the necessary authentication details from the *patient_ids*.*json* file to initialize tracking for that patient. Once the account becomes active on any device, the script automatically sends an email notification to a designated account owned by our team indicating that a session has started and begins logging playback and device information in real time. More specifically, the script tracks and stores the following information: patient ID, session start time in EDT, artist name, album name, track name, track’s original duration, starting position of the track in seconds (whether it starts from the beginning or not), session start and end timestamps, track listening duration, total track listening duration (accumulated if the track is repeated), track end reason (end of song, repeated, skipped, seeked forward/backward, paused, or Spotify closed), and the device name that the account was used on (e.g., iPad, MacBook). We use Python’s datetime library to track time and calculate the track start position, track listening duration, total track listening duration, and session duration. This will enable precise calculation of treatment dose.

When the user closes the Spotify app or remains in a paused state for more than five minutes, the session is marked as finalized. Then the script sends an email notification with the total session duration, saves the information as a .csv file in the designated folder for that patient, and uploads it to an encrypted, password protected, Google Drive account. The code for this software can be found here: https://github.com/intentional-music-listening/SpotifyTrackerOpen

#### Acoustic Fingerprinting

With the exact list of music that patients listened to, we will use MIR methods and open-source software (e.g., *librosa*; McFee et al., 2015) to analyze the acoustic and musical features of the music participants listen to daily. Different measures will be extracted, based on previous research showing that specific acoustic properties of music (e.g., tonality, rhythm, timbre, loudness, surprise, amount of vocal vs. instrumental music; Carone and Ripollés, 2024; Groves et al., 2025; Sihvonen et al., 2020, Sihvonen et al., 2021a, Sihvonen et al., 2021b; Salakka et al., 2022; Abrams et al., 2022; Skerritt-Davis & Elhilali, 2018) can affect emotional and reward related responses to music and modulate mental health and cognition (Ding et al., 2023; Parada-Cabaleiro et al., 2022; Bowling, 2023).

For control participants, acoustic fingerprint measures will be extracted from the audiobooks. Using an Audio-to-Text transcriber, we will convert the exact audiobook section each participant listened to into text. From these transcripts, we will compute lexical (e.g., average word frequency, average number of morphemes per word), semantic (e.g., average number of semantic neighbors per word), and syntactic (e.g., number of phrases and nested structures) complexity measures, among others. These analyses will be performed using open-source toolboxes such as the Stanford Parser (Klein & Manning, 2003; Yap et al., 2011).

#### Post-intervention evaluation session

Following the 4-week intervention period, participants will complete a post-intervention evaluation session identical to the pre-intervention session. This assessment will include clinical outcomes related to mental health and cognition, neural and physiological biomarkers (functional and structural MRI, physiological data), and burden ratings.

#### Follow-up evaluation session

Three months after completing the intervention, participants will complete a final follow-up evaluation session. This session will assess only behavioral clinical outcomes related to mental health and cognition; no biomarkers will be collected.

### Outcome measures

This study is a clinical trial assessing the feasibility of an IML, and therefore the primary outcomes will focus on feasibility metrics. Primary outcomes will include retention and adherence, feasibility, acceptability, fidelity, and burden. Secondary outcomes will include recruitment and randomization. Table 2 provides a detailed description of each outcome along with predefined benchmarks for success.

**Table 2.** Outcome measures and benchmarks for success.

| Outcome | Description | Benchmark for success |
| --- | --- | --- |
| Recruitment | Number of enrolled participants per month | At least 2 per month |
| Randomization | Proportion of screened patients who are enrolled<br>Proportion of enrolled patients who complete the baseline and pre-intervention evaluations and attend at least one IML/Audiobook session | At least 30%<br>At least 80% |
| Retention and Adherence | Proportion of enrolled and randomized participants who complete the IML/Audiobook sessions | At least 80% of randomized patients complete at least 80% of the IML/Audiobook sessions |
| Feasibility | Proportion of listening sessions considered fully feasible; the team member making the phone call will rate the session as “not feasible” , “partially feasible” or “fully feasible” | At least 80% of all sessions considered as fully feasible |
| Fidelity | Proportion of listening sessions delivered according to the protocol; video calls with patients will be recorded and a team member who was not present during the call will score the session as “delivered according to protocol” or “not delivered according to protocol” | At least 80% of all sessions delivered according to the protocol |
| Burden | Proportion of planned evaluations that are completed at each time point (baseline, pre-intervention, post-intervention, follow-up)<br>Burden rating: patients will rate each evaluation as “not burdensome” , “partially burdensome” , or “very burdensome” | 80% of patients complete the evaluations at each time point<br>At least 80% of patients rate evaluations at each time point as not burdensome |
| Patient acceptability | Acceptability ratings: patients will rate each listening session as “not acceptable” , “partially acceptable” or “fully acceptable” . | 80% of listening sessions rated as fully acceptable |

### Statistical analysis

The primary and secondary outcomes of intervention feasibility will be reported using percentages (see Table 2, benchmarks for success). One-sided exact binomial tests and proportional z-tests will be used to assess whether the outcomes have success rates above the benchmarks. Success rates will be compared between the treatment group and control group using Chi-squared tests or Fisher’s Exact Tests if non-normality presents. We will identify potential factors that are associated with the feasibility outcomes, which will help refine the design of a large-scale clinical trial.

### Patient and public involvement

The proposed protocol has been informed, revised, and refined in consultation with stroke patients.

### Ethics and dissemination

The study was approved by New York University’s Institutional Review Board (FY2024-8826) and was registered on ClinicalTrials.gov (NCT07127159). The study follows the recommendations of the NIH Music-Based Intervention Toolkit (Edwards et al., 2023) and of SPIRIT (Standard Protocol Items: Recommendations for Interventional Trials; Chan et al., 2013). All participants will provide written informed consent prior to enrollment. Compensation will be provided for each completed evaluation session, and transportation costs will be covered as needed. As part of ethical approval, this protocol includes a Data and Safety Monitoring Plan (DSMP) and an independent monitoring committee (IMC). The IMC is composed of three members who are not involved in this research project and operate independently of the principal investigators. They have not collaborated or co-published with the principal investigators in the past three years. Each member brings relevant expertise that qualifies them to review the data generated by the study. The IMC will receive one report annually and will provide feedback and recommendations based on their review. Monitoring of study data will occur at multiple levels and frequencies. The Principal Investigator (Ripollés) will conduct monthly reviews of subject accrual, participant status, data entry quality, adverse events (AEs), and unanticipated problems. Ripollés will also monitor serious adverse events (SAEs) per occurrence. The IMC will review subject accrual, participant status, and AEs annually, and will also review SAEs per occurrence (if unexpected and related) or annually (if expected or unrelated). Additionally, the National Center for Complementary and Integrative Health (NCCIH) of the National Institutes of Health (USA) will review AEs annually and SAEs per occurrence (if unexpected and related) or annually (if expected or unrelated). The Institutional Review Board will review unanticipated problems according to policy. If the number of overall SAEs exceeds 33% (i.e., 1 in 3) of recruited patients the study will be suspended, and an ad-hoc safety review will be completed. All protocol amendments, except for minor administrative changes, will be submitted prospectively to the NCCIH unless immediate changes are needed to protect participant safety, rights, or welfare. Proposed amendments will first be reviewed and approved by the IMC, and IRB approval will be sought only after approval has been obtained by the NCCIH.

A special provision has been established under the DSMP to address potential mental health crises. The Beck Depression Inventory, administered at all in-person evaluations, includes a question assessing suicidal ideation. Responses to this item will be reviewed immediately. If a participant selects a score of 1, 2, or 3, a predefined protocol will be activated. Participants assessed to be at acute risk will be referred for emergency psychiatric care, either through a hospital emergency department or via 911. Those experiencing significant but non-acute distress will be provided with appropriate clinical resources, including a list of wellness tools, self-care apps, and mental health clinics in New York City that accept various insurance plans (see Supplementary Materials).

All data will be keyed with an anonymous code. All personal information will be treated as confidential; only authorized personnel will have access to this data, and it will not be given to third parties without the subject’s explicit consent. All personally identifiable information will be stored in a secure, encrypted server maintained explicitly for the storage of highly sensitive data, including protected health information. Only the research team will have access to this encrypted data. Experimental results, devoid of any personal information that could identify them, will be maintained in the principal investigator’s files, and eventually released as an open-source database. Results will be reported in peer-reviewed journals. Study completion is expected by Fall 2026.

## Discussion

This paper presents the protocol for a feasibility clinical trial evaluating a remote IML intervention for individuals with chronic stroke. The study includes an objective measure of treatment dose and aims to support mental health recovery in this population. For the feasibility clinical trial, we expect to meet the benchmarks specified in Table 2. In a future efficacy trial informed by this protocol, we expect that participants in the IML group will show improvements in mental health and cognition, accompanied by neuroplastic changes that reflect and support these behavioral gains, compared to the audiobook control group. This hypothesis is grounded in prior research showing that music listening engages multiple brain systems related to the mechanistic pathways (motor, sensory, cognition, and social) through which an enriched environment supports brain plasticity and recovery (Zatorre et al., 2007; Rodriguez-Fornells et al., 2012; Koelsch, 2014; Mas-Herrero et al., 2021; Janata et al., 2002). Importantly, music listening has been shown to activate key brain regions involved in reward, dopaminergic processing, and motivation (Ferreri et al., 2019; Mas-Herrero et al., 2021; Koelsch, 2014). This is particularly relevant given prior research suggesting that the mental health benefits of music are closely linked to reward-related mechanisms (Mas-Herrero et al., 2023). We also hypothesize that improvements in mental health and cognitive functioning will be directly related to the level of treatment dose.

The intervention is guided by the NIH Music-Based Intervention Toolkit (Edwards et al., 2023) and is designed to be low-cost, accessible, scalable, and highly acceptable to patients. Music listening interventions can also optimize the use of sedentary time spent by stroke survivors, as patients can engage in the intervention from home, regardless of motor limitations or access to specialized stroke and/or rehabilitation units (Baylan et al., 2016; Oermann and Riina, 2021). Accurately measuring treatment dose is essential for optimizing rehabilitation outcomes (Schambra et al., 2019). For example, high-intensity, repetitive motor activity is a strong predictor of recovery in stroke rehabilitation (Hatem et al., 2016). An advantage of our approach is our objective quantification of music treatment dose, which has not been quantified in previous IML studies in stroke populations. In the context of IML, dose refers to not only the duration of the music but also to the specific acoustic and musical features of the selected music. These features may differentially impact mental health (Bowling et al., 2023; Orpella et al., 2025). This protocol introduces the use of real-time tracking to objectively measure listening behavior. This approach allows to quantify treatment dose not only by duration but also by analyzing the content of the music. By assessing the relationship between acoustic/musical features and changes in mental health, we can identify which aspects of music may be most therapeutically relevant. This methodology will allow future large-scale, sufficiently powered, clinical trials to assess what amount of IML and which characteristics of the music listened to specifically modulate mental health, cognitive recovery, and brain plasticity.

A strength of this study is access to a large, diverse, and well-characterized population of chronic stroke patients from the greater New York City area. Another strength of the study is the use of multimodal methods to assess intervention effects across behavioral, physiological, and neuroimaging (MRI) domains, providing a more comprehensive understanding of the intervention’s impact. However, the study has limitations. It is not powered to detect clinical changes in mental health, cognition, or neuroplasticity, as its primary goal is to assess feasibility. Additionally, as a single-center study, generalizability to other clinical settings or populations may be limited.Regardless of the outcomes of the proposed work, we believe this study will help advance the field of music-based interventions at the theoretical, experimental, and clinical level. The IML intervention tested here may also be adaptable to other aging-related neurological conditions, such as Alzheimer’s disease and other dementias, broadening its potential public health impact.

## Acknowledgments

We would like to thank Dr. Lucia Vaquero for her help in developing the mental health resources document.

## Data availability statement

All data is included in the main manuscript and supplementary materials

## Author Contributions

According to the CRediT (Contributor Roles Taxonomy), author contributions to this study were as follows: Conceptualization was led by Ripollés, Fuentes, and Schambra. Methodology was developed collaboratively by Provias, Ripollés, Fuentes, Schambra, Kucukoglu, Robinson, Yandun-Oyola, He, Palumbo, Malgaroli, Sihvonen, and Shi. Software development was carried out by Fuentes, Kucukoglu, Provias, and Palumbo. Validation and formal analysis were conducted by Provias, Robinson, Yandun-Oyola, Ripollés, Fuentes, and Kucukoglu. Investigation efforts were led by Provias, Robinson, Yandun-Oyola, and Kucukoglu. Resources were provided by Fuentes, Ripollés, Malgaroli, and Schambra. Data curation was managed by Provias, Robinson, Yandun-Oyola, Kucukoglu, Ripollés, Fuentes, and Schambra. The original draft was written by Provias, Ripollés, Fuentes, Kucukoglu, Robinson, and Yandun-Oyola, with all authors contributing to review and editing. Visualization was prepared by Provias and Ripollés. Supervision and project administration were led by Ripollés and Fuentes. Funding acquisition was carried out by Ripollés, Fuentes, Schambra, Sihvonen, Malgaroli, and Shi.

