## Supplemental Materials for "Feasibility Trial Protocol for a Remote Intentional Music Listening Intervention to Support Mental Health in Individuals with Chronic Stroke"

### Supplementary Materials

List of Supplementary Materials

1. Demographics questionnaire
2. MRI screening questionnaire
3. Musical preferences questionnaire
4. Audiobook preferences questionnaire
5. Control measures intentional music listening group (mEMA app)
6. Control measures audiobook control group (mEMA app)
7. Mental health resources document

**DEMOGRAPHICS QUESTIONNAIRE**

Subject code: _______________

Participant Name _____________________________________________

Email Address _____________________________________________

Age (in years) _____________________________________________

**Gender**

- ☐ Male
- ☐ Female
- ☐ Non-binary/third gender
- ☐ Prefer not to say

**Race & Ethnicity**

How would you describe yourself? Select all that apply:

- ☐ American Indian or Alaskan Native
- ☐ Asian
- ☐ Black or African American
- ☐ Native Hawaiian or Other Pacific Islander
- ☐ White
- ☐ Other, please specify _______________

Are you of Hispanic, Latino/a, or Spanish origin?

- ☐ Yes
- ☐ No

**Handedness**

Which is your dominant hand?

- ☐ Right
- ☐ Left

**Languages**

Which languages do you speak? Please list:

_____________________________________________________________

_____________________________________________________________

_____________________________________________________________

_____________________________________________________________

**Medications**
Please list all medications you are currently taking, with their dose:

________________________________________________________________
________________________________________________________________
________________________________________________________________
________________________________________________________________

**Additional Health Activities**
Please list any complimentary health activities that you regularly practice
and specify how often you do them. Examples: yoga, acupuncture, meditation, exercise

________________________________________________________________
________________________________________________________________
________________________________________________________________
________________________________________________________________

**Occupational Status**
Please select one of the following:

- ☐ Still at school
- ☐ In University
- ☐ Full-time employment
- ☐ Part-time employment
- ☐ Self-employed
- ☐ Homemaker/Full-time parent
- ☐ Unemployed
- ☐ Retired

**Education**
What is the highest educational qualification you have attained?

- ☐ Did not complete any school qualification
- ☐ Completed first school qualification (GCSE/Junior High School)
- ☐ Completed second school qualification (A Levels/High School)
- ☐ Undergraduate degree or professional qualification
- ☐ Postgraduate degree
- ☐ I am still in education

If you are still in education, what is the highest qualification you expect to obtain?

- ☐ first school qualification (GCSE/Junior High School)
- ☐ second school qualification (A Levels/High School)
- ☐ Undergraduate degree or professional qualification
- ☐ Postgraduate degree
- ☐ Not applicable

NYU | ARTS & SCIENCE

Center for Brain Imaging

### Subject Screening Form

**Warning!** Certain implants, devices or objects may be hazardous to you and or may interfere with the Magnetic Resonance (MR) system. **Do Not Enter** the MRI control room or MR environment if you have any question or concern regarding an implant, device or object. Consult with the Researcher or MRI Technologist before entering the MR system room. **The MR System Magnet Is Always On.**

**Note**: You will be advised or required to wear earplugs or other hearing protection during the MRI to prevent possible problems or hazards related to acoustic noise. Please consult with the Researcher or MRI Technologist if you have any questions.

Date: ______________

Last Name: ____________________________________ First Name: ____________________________

Age: _______ Weight: _________ Gender _______ Race/Ethnicity: (optional) _____________________

Principal Investigator: _____________________________________ Study ID: __________________

| **Please indicate if you have any of the following:** | **Yes** | **No** |
|---|---|---|
| o History of head trauma | ___ | ___ |
| o Surgical aneurysm clips | ___ | ___ |
| o Cardiac pacemaker (even if removed) | ___ | ___ |
| o Prosthetic heart valve | ___ | ___ |
| o Neurostimulator | ___ | ___ |
| o Transdermal patch (nicotine, nitroglycerin, hormone) | ___ | ___ |
| o Implanted pumps | ___ | ___ |
| o Cochlear implants | ___ | ___ |
| o Metal rods, plates, screws | ___ | ___ |
| o Previous surgery | ___ | ___ |
| o IUD | ___ | ___ |
| o Hearing Aid | ___ | ___ |
| o Dentures, braces, or non-removable retainer | ___ | ___ |
| o Hair Extensions | ___ | ___ |
| o History of injury to eye involving metal | ___ | ___ |
| o Any metal in the body – shrapnel, bullets or buckshot | ___ | ___ |
| **Please answer the following questions:** | | |
| o Do you typically experience Claustrophobia? | ___ | ___ |
| o Are you under the care of an epileptologist? | ___ | ___ |
| o Is there any chance you might be pregnant? | ___ | ___ |
| o Are you wearing colored contact lenses? | ___ | ___ |
| o Do you have any tattoos on your face (e.g., eyeliner tattoo) or neck, back, or groin; tattoos that cover more than 5% of your body, and/or a single tattoo that is larger than 20cm/8in? | ___ | ___ |
| o Have you ever worked as welder or metal worker? | ___ | ___ |

If you answered "**YES**" to any of the items above, please explain:

---

I have received a copy of the informed consent document for this study (initial here): __________

Signature: ___________________________________________________________ Date: ____________

Witness: _____________________________________________________________ Date: ____________

***DO NOT ENTER THE SCAN ROOM WITH ANY OF THESE ITEMS***

| | | | | |
|---|---|---|---|---|
| Credit/Bank Cards | Wallet/Money Clips | Underwire Bra | Pens/Pencils | Wigs/Hairpiece |
| Body Piercing | Hairpins/Barrettes | Safety Pins | Glasses | Coins |
| Watch | Keys | Pocket Knife | Jewelry | Belts/Buckles |

**MUSIC PREFERENCES**

Subject code: _______________

**Genre Preference**

What type of music do you like to listen to? Check all that apply:

- ☐ Classical
- ☐ Pop
- ☐ Jazz
- ☐ Electronic
- ☐ Hip-hop
- ☐ Soul
- ☐ Country
- ☐ If other, please specify _______________

**Mood**

How do you prefer the mood of your music? Check all that apply:

- ☐ Uplifting / Happy
- ☐ Calm / Relaxing
- ☐ Energentic / Exciting
- ☐ Melancholic / Thoughtful
- ☐ No preference

**Bands / Artists**

Please provide us with a list of 5-10 artists that you enjoy listening to

____________________________________________________________

____________________________________________________________

____________________________________________________________

____________________________________________________________

____________________________________________________________

____________________________________________________________

____________________________________________________________

____________________________________________________________

____________________________________________________________

____________________________________________________________

**Songs**

Please provide a list of 6 songs that give you musical chills and are particularly pleasurable to you.

____________________________________________________________

____________________________________________________________

____________________________________________________________

____________________________________________________________

**AUDIOBOOK PREFERENCES**

Subject code: _______________

**Genre Preference**

What types of books do you enjoy? Check all that apply:

- ☐ Fiction
- ☐ Non-fiction
- ☐ Mystery / Thriller
- ☐ Romance
- ☐ Science Fiction / Fantasy
- ☐ Biography / Memoir
- ☐ Self-help / Psychology
- ☐ History / Politics
- ☐ Science / Nature
- ☐ Poetry
- ☐ If other, please specify: _______________

**Mood**

What kind of tone or mood do you prefer in books? Check all that apply:

- ☐ Light-hearted / Funny
- ☐ Thought-provoking / Reflective
- ☐ Suspenseful / Intense
- ☐ Uplifting / Motivational
- ☐ Dark / Emotional
- ☐ No preference

**Authors**
Please list some of your favourite authors:

____________________________________________________________
____________________________________________________________
____________________________________________________________
____________________________________________________________

**Books**
Please list 1-3 books you would like to listen to:

____________________________________________________________
____________________________________________________________

#### CONTROL MEASURES FOR INTENTIONAL MUSIC LISTENING GROUP

1. **On a scale from 1 to 5, how COGNITIVELY active have you been today?**
   Examples: reading, solving puzzles, doing Sudoku, writing

   1 2 3 4 5

2. **On a scale from 1 to 5, how PHYSICALLY active have you been today?**
   Examples: walking, running, exercising, physical work, cleaning

   1 2 3 4 5

3. **On a scale from 1 to 5, how SOCIALLY active have you been today?**
   Examples: talking to friends or family, social gatherings, volunteering, community activities

   1 2 3 4 5

4. **How many minutes of music have you been exposed to OUTSIDE of today's music listening session?** Examples: listening to music on the radio, playing an instrument, or going to a concert

   - ☐ None
   - ☐ Less than 30 minutes
   - ☐ Between 30 and 60 minutes
   - ☐ 1-2 hours
   - ☐ More than 2 hours

5. **On a scale of 1-5, when you were listening to music outside of the listening today, how attentive were you to it?**

   1 2 3 4 5

CONTROL MEASURES FOR AUDIOBOOK CONTROL GROUP

1. **On a scale from 1 to 5, how COGNITIVELY active have you been today?**
   Examples: reading, solving puzzles, doing Sudoku, writing

   1 2 3 4 5

2. **On a scale from 1 to 5, how PHYSICALLY active have you been today?**
   Examples: walking, running, exercising, physical work, cleaning

   1 2 3 4 5

3. **On a scale from 1 to 5, how SOCIALLY active have you been today?**
   Examples: talking to friends or family, social gatherings, volunteering, community activities

   1 2 3 4 5

4. **How much time have you spent listening to the audiobook outside of today's session?**

   - ☐ None
   - ☐ Less than 30 minutes
   - ☐ Between 30 and 60 minutes
   - ☐ 1-2 hours
   - ☐ More than 2 hours

5. **On a scale of 1-5, when you were listening to the audiobook outside of the listening today, how attentive were you to it?**

   1 2 3 4 5

6. **How many minutes of music have you been exposed to today?** Examples: listening to music on the radio, playing an instrument, or going to a concert

   - ☐ None
   - ☐ Less than 30 minutes
   - ☐ Between 30 and 60 minutes
   - ☐ 1-2 hours
   - ☐ More than 2 hours

### Mental Health Resources Document

#### 1. Apps and Sites

Here are some suggested apps you may find helpful, with tips and guided practices to improve your motivation, self-discovery, and mental health, as well as community to exchange ideas and receive support or relatable testimonials:

- ☐ Happify
- ☐ HeadSpace
- ☐ Sanvello
- ☐ Mindset

The Anxiety and Depression Association of America (ADAA) website also offers a more extensive list of apps, as well as reviews by mental health professionals: https://www.adaa.org/finding-help/mobile-apps

#### 2. Exercise

Some activities that can be beneficial to your mental and physical well-being include:

- ☐ Yoga. DoYogaWithMe.com offers free yoga video classes for all levels
- ☐ Sports, running, walking
- ☐ Any level of physical activity can be helpful, from a brisk 30-minute walk through the park to playing a recreational sport or going for a jog.

Interested in learning more? Here are some books on the topic:

- *Exercise for Mood and Anxiety: Proven Strategies for Overcoming Depression and Enhancing Well-Being* by Michael Otto

#### 3. Mindfulness and Meditation

Even practicing 5 minutes of mindfulness or meditation a day can help you relax. The Greater Good Science Center at UC Berkeley provides short and accessible guided mindfulness meditation practices you can do on your own. These can be found online at: http://ggia.berkeley.edu/.

Here are some highlighted practices:

- ☐ Mindful breathing – this exercise takes 15 minutes and includes focusing your attention on your breath.
- ☐ Self-compassion break – a 5-minute exercise designed to help after a stressful situation.
- ☐ Body scan – 30 minutes of guided full-body meditation.

Some recommended books on mindfulness:

- *The Mindfulness Revolution* by Barry Boyce
- *Fully Present* by Susan Smalley and Diana Winston
- *The Happiness Project* by Gretchen Rubin

For more helpful mindfulness and meditation resources such as videos, published studies, organizations, and books, visit the Greater Good website: http://greatergood.berkeley.edu/

#### 4. Nutrition, Sleep, and Self-Care

Caring for your body is equally as important as caring for your mind. This includes eating well and getting enough sleep (though we all know how difficult that can be!).

The American Heart Association website offers advice on how to make healthier food choices and also provides recipes and a nutrition guide – https://www.heart.org/en/healthy-living/healthy-eating

The American Academy of Sleep Medicine offers guidelines on how get a good night's rest: https://sleepeducation.org/healthy-sleep/healthy-sleep-habits/

#### 5. Books about mental health

Listed below are books that address mental health issues written for the reader who may want to understand more about depression, anxiety, and stress, and about how to cope with these feelings:

- *Mind Over Mood* by Dennis Greenberger and Christine Padesky
- *Retrain your Brain* by Seth J. Gillihan
- *Full Catastrophe Living* by Jon Kabat-Zinn
- *The Anxiety and Phobia Workbook* by Edmund J. Bourne

#### 6. Suicide Prevention

In light of growing public awareness of this issue, the Anxiety and Depression Association of America (ADAA) website has articles dedicated to suicide prevention and resources.

- [ ] https://adaa.org/understanding-anxiety/suicide
- [ ] https://adaa.org/understanding-anxiety/suicide-resources

The National Suicide Prevention Lifeline is a 24-hour service dedicated to offer help for both people in distress as well as for professionals and loved ones who may be concerned.

- [ ] https://suicidepreventionlifeline.org/

#### 7. Referrals

The clinics listed below, by location, accept most insurance providers.

##### MANHATTAN

NewYork-Presbyterian/Weill Cornell Medical Center - Payne Whitney Clinic. 525 East 68th Street, New York, NY 10065. Children, adolescents, and adults receive the most current therapeutic and pharmacologic care available to treat mood, anxiety, attention deficit, personality, and other disorders. At either facility, patients can receive psychiatric evaluations by a qualified mental health therapist or psychiatric doctor. For an intake call: (646) 962-2820.

NYC Health Hospital: Bellevue-The Bellevue Outpatient Psychiatry Clinic. First Avenue and 27th Street, Ambulatory Care Building, Ground Floor RM-G1027. Clinic Hours: Monday - Friday, 8:30 am - 4:30. To make or change an appointment, call (212) 562-5710. Check for insurance accepted here.

Lenox Hill Hospital-Outpatient Center for Mental Health. 210 E. 64th St. NYC, NY 10065. (212) 434- 3365. This clinic provides individual, group, family therapy, neuropsychological testing, and medication management. Accepts most forms of health insurance.

Mount Sinai West. 1000 Tenth Avenue New York, NY 10019. Phone: 212-523-4000. Mount Sinai West, provides comprehensive, high-quality psychiatry and behavioral health services.

Outpatient Mental Health Services at Mount Sinai Morningside Hospital. 1111 Amsterdam Avenue, New York, NY 10025. For questions or to schedule an initial evaluation, please call 332-243-0080. The phone line is open from 8:30 am to 4 pm Monday through Friday

Martha Stewart Center for Living at Mount Sinai Hospital. 17 East 102nd Street, 4th Floor, Area C, New York, NY 10029. Phone: 212-659-8552. Specialized in the evaluation, diagnosis, and treatment of those individuals who are over 60 years of age with various mood, anxiety, and psychotic disorders, as well as dementia.

Mount Sinai-Behavioral Health Center. 45 Rivington Street, New York, NY 10002. Phone: 332-243-1600. Specialized in the evaluation, diagnosis, and treatment of those individuals who are over 60 years of age with various mood, anxiety, and psychotic disorders, as well as dementia.

NewYork-Presbyterian/Columbia Psychiatry. 630 West 168th Street New York, NY 10032. Phone: 212-305-6001. This center provides individual and group therapy, family therapy, electroconvulsive therapy or medication (as needed), and social support services.

NYC Health Hospital:Harlem. 506 Lenox Avenue, New York, NY 10037. General Information: 212-939-1000. Appointments: 844-692-4692. This hospital includes a Comprehensive Psychiatric Emergency Program (CPEP) to treat patients in mental and emotional crisis situations (phone: 212-939-3343). It also includes an Adult Outpatient Clinic (phone: 212-939-8491). Check for insurance accepted here.

#### BRONX

Montefiore Behavioral Health Center. 2527 Glebe Avenue, Bronx NY, 10461. Please call 718-904-4476 or for an appointment. Provides individual psychotherapy, group psychotherapy and medication management services, and primary care services. Check insurance accepted here.

Moses Adult Outpatient Psychiatry Program. 111 East 210th Street, Klau 1, Bronx NY, 10467. Phone: 718-920-4295. Services offered: individual psychotherapy, group psychotherapy and medication management services, including long acting injectables. Check insurance accepted here.

Bronx Psychiatric Center. 1500 Waters Place, Bronx, NY 10461. Phone: (718) 931-0600. Specializes in treatment of people with severe and complex mental illness who need care over time. Operates in collaboration with County Departments of Mental Health.

NYC Health Hospital: Jacobi. 1400 Pelham Parkway South, Bronx, NY 1046. General Information: 718-918-5000. Appointments: 844-692-4692. The Behavioral Health Service at Jacobi offers a comprehensive continuum of consultative, inpatient, ambulatory, and emergency services for people with mental health and substance abuse disorders. It includes the Comprehensive Psychiatric Emergency Program (CPEP). CPEP is a hospital-based psychiatric emergency program which provides a primary entry point to the program for individuals who may be mentally ill to receive emergency observation, evaluation, care, and treatment in a safe and comfortable environment. CPEP is open 24/7 and its phone is 718-918-4850. Check for insurance accepted here.

NYC Health Hospital: Lincoln. 234 East 149th Street, Bronx, NY 1045. General Information: 718-579-5000. Appointments: 844-692-4692. Lincoln's Department of Behavioral Health provides comprehensive mental health services to individuals and families. A continuum of services is offered that includes inpatient as well as outpatient psychiatric care. Check for insurance accepted here.

NYC Health Hospital: North Central Bronx. 3424 Kossuth Avenue, Bronx, NY 10467. General Information: 718-519-5000. Appointments: 844-692-4692. The Behavioral Health Services department includes inpatient psychiatric care, addiction treatment, and outpatient emotional and mental health services for children, adolescents, adults, and seniors. This hospital includes a Psychiatric Emergency Department (1st Floor, 1D-01; open 24/7; phone: 718-519-3030) that provides timely, high quality, effective treatment in a safe and caring environment. Emergency care is provided by a multidisciplinary clinical staff in a private and confidential setting. Services include psychiatric diagnostic evaluation, crisis intervention, psychopharmacologic and psychotherapeutic interventions, medical assessment, discharge planning, and linkage to community resources. The hospital also includes a Geriatric Psychiatry Inpatient Unit (12th Floor, 12A; phone: 718-519-2072) that is dedicated exclusively to providing specialized mental health services for the elderly. Check for insurance accepted here.

#### BROOKLYN

NYC Health Hospital: Kings County. 451 Clarkson Avenue, Brooklyn, New York 11203. General Information: 718-245-313. Appointments: 844-692-4692. The Behavioral Health Services (BHS) center for adults is comprised of: Mental Health Outpatient clinic, Esketamine Clinic, Primary Care Clinic, Partial Hospitalization Program, Intensive Outpatient Program, the Family Justice Center (FJC) for domestic violence care, Kings on Track (KOT), and Kings Early Episode Program (KEEP) for our newly diagnosed patients. Services are available by directly walking-in to the R-Building; Monday-Friday from 8:30-4. To reach the BHS directly call 718-245-2700/2704. The BHS also contains a Comprehensive Psychiatric Emergency Program (CPEP), that includes a 24/7 Psychiatric Emergency Room; Extended Observation Unit (6 beds) and Mobile Crisis Team. Entrance to CPEP is on Winthrop Street at New York Avenue. Check for insurance accepted here.

NYC Health Hospital: South Brooklyn Health. 2601 Ocean Parkway, Brooklyn, NY 11235. Appointments: 844-692-4692. The behavioral health center at South Brooklyn Health provides both inpatient and outpatient treatment for behavioral health needs. The center includes a Psychiatric Emergency Service that provides the assessment, intervention, treatment, and referral services for mental health crises. The center also includes Outpatient Services for adults presenting with symptoms of psychiatric and co-morbid substance use disorders. For more information about behavioral health services or to schedule an appointment, please call 844-692-4692. Check for insurance accepted here.

NYC Health Hospital: Woodhull. 760 Broadway, Brooklyn, New York 11206. General Information: 718-963-8000. Appointments: 844-692-4692. The Department of Behavioral Health at Woodhull provides comprehensive mental health services to individuals and families. A continuum of services is offered that includes inpatient as well as outpatient psychiatric care. The department is staffed by a multidisciplinary team of psychiatrists, licensed clinical psychologists, mental health therapists, nurses, and social workers who work in collaboration to develop recovery plans tailored to meet the clients' individual needs. Check for insurance accepted here.

Sunset Terrace Family Health Center at NYU Langone. 514 49th St, Brooklyn, NY 11220 General Information: 718-437-5210. The Behavioral Health Program - Family Health Centers at NYU Langone provides treatment and support for people and families who are experiencing mental health disorders across all age ranges. Care is provided for people experiencing depression, mood and anxiety disorders, ADHD, in adults and children, insomnia, and schizophrenia and psychotic disorders, as well as other mental health conditions. Check for insurance accepted here.

Maimonides Medical Center: 4802 Tenth Avenue, Brooklyn, NY 11219. Adult Outpatient Services: 718-283-7800. The Behavioral Health department offers comprehensive assessment, treatment planning, and therapeutic services to meet the mental health and developmental needs of children, adults, and families in Brooklyn and beyond. The department includes a Consultation and Emergency Psychiatry Service that provides psychiatric consultations for adults (phone: 718-283-7841). Check for insurance accepted here.

Brooklyn Connected Certified Community Behavioral Health Clinic (CCHBC). 2020 Coney Island Ave., Brooklyn, NY 11223.. Phone: 844-663-2255. CCBHCs provides multiple services to aid people with any mental health or substance use problems. CCBHCs aims to help individuals regardless of a person's ability to pay or residence– this includes the underserved, those with little to no income, those that are insured as well as those who are uninsured or on Medicaid.

#### QUEENS

NYC Health Hospital: Elmhurst. 79-01 Broadway Elmhurst, New York 11373. General Information: 718-334-4000. Appointments: 844-692-4692. Services provided encompass many levels, including: emergency services, acute care hospitalization, ambulatory behavioral health services, and community oriented services. For specific information on behavioral and mental health services at NYC Health Elmhurst, please call 718.334.4000. Check for insurance accepted here.

NYC Health Hospital: Queens. 82-68 164th Street Queens, New York 11432. General Information: 718-883-3000. Appointments: 844-692-4692. The Psychiatry Department at NYC Health Queens includes, among others: a Comprehensive Psychiatric Emergency Services Program (CPEP), offering 24-hour services (phone: 718-883-3575); a Mobile Crisis Unit, providing a team consisting of a psychiatrist, psychologist and social worker who can call or visit the home or workplace of a patient and make an on-site assessment (phone: 718-883-4070); adult outpatient services (phone: 718-883-2725); and an Assertive Community Treatment Program (ACT) that provided delivers treatment to patients with persistent mental illness who are unable or unwilling to participate in traditional models of care in the community (phone: 718-883-6561). Check for insurance accepted here.

North Shore- Long Island Jewish Child and Adolescent Psychiatry: Cohen Children's Medical Center: 75-59 63 Street Glen Oak, New York 11004. (718) 470-3500. This clinic offers individual, group, and family therapy, as well as medication management.

St. John's Center for Psychological Services: 152-11 Union Turnpike, Flushing, NY 11367. Phone: 718-990-1900. This clinic provides family therapy, individual therapy, and psychoeducational testing. They do not take insurance but the fee for services is based on a sliding scale based on household income.

#### STATEN ISLAND

NYC Gotham Health: Staten Island. 165 Vanderbilt Avenue, Staten Island, NY 10304. Appointments: 844-692-4692. Hours of Operation: Mon., Wed., Thu., Fri.: 8:00 a.m. 5:00 p.m; Tue.: 8:00 a.m. 7:00 p.m.; Sat.: 8:30 a.m. 4:30 p.m.; Sun.: Closed. This clinic provides the full spectrum of care for children and adults in Staten Island, including mental health services. Check for insurance accepted here.
